# Role of Whole Body CT (WBCT) in Trauma Patients “Life Saver or Needless Radiation Exposure?”

**DOI:** 10.1101/2020.06.20.20136267

**Authors:** Latifa Alkandari, Mahdy A Abass, Michael Masoomi, Shreeram Kannan, Samuel D Ashebu, Hagrassy Abdulla

## Abstract

**Background:** Whole-body computed tomography (WBCT) is used indiscriminately in trauma cases, just on the suspicion of them being polytrauma cases. A good clinical examination done pre-emptively could prevent the need for this investigation and its undesired radiation effects. The use of WBCT was assessed in our busy hospital to determine whether there has been an overuse of the WBCT and also to estimate the true incident of clinical injuries.

**Methods:** Retrospective database analysis of 546 WBCT polytrauma cases for the period of April to October 2018 was performed. All the trauma patients were initially managed and proceed for WBCT according to the American College of Radiology (Major Trauma). We recorded age, gender, mechanism of injury, clinical requests, WBCT findings in regions of the cervical spine, thoracolumbar spine, chest, abdomen and pelvis skeleton injuries (as per our institutional reporting protocol), as well as DLP for each patient scanned. We compared pre-test clinical requests stating the mechanism of injury and clinical query with WBCT findings and categorized the radiological findings, initially into negative and positive findings. The positive findings were further classified into the major and minor injuries. The total numbers in each radiological finding were calculated and inferences discussed.

**Results:** On analysis of data, we found that 462 patients had been referred due to RTA (84.6%), 47 patients due to FFH (8.6%), ten patients due to blunt trauma (1.8%), 8 patient due to assault (1.5%), while the 5 patients had stable wound (0.09%), five patients had injury due to the fall of a heavy object (0.09%), 3 patients had a buggy injury (0.05%), 3 patients had injury due to fallen on the back (0.05%) and 3 more patients had injury due to other traumas including blast injury, suicide and other injury. We noticed RTA with an 84.7 % score was the most common indication for WBCT referral. Out of 546 cases, 414 patients (75.8 %) were normal (negative finding), where 132 patients (24.2 %) had positive trauma related radiological finding of which 54 patients were found to have a major injury (9.9%). Fractures were scored the highest, 75.6% of all positive finding traumas.

**Conclusions:** This study re-emphasizes the significance of exercising a good clinical examination in the era of evidence based medicine, which would reduce the high number of unnecessary high dose WBCT, as 462 scans with no positive findings on radiological examinations were nearly normal and only 54 cases (9.9%) had major injuries.

## INTRODUCTION

The use of whole-body computed tomography (WBCT) is advocated for rapid and comprehensive objective diagnosis of serious injuries. The proponents of WBCT justify its use due to its potential benefit in detecting unsuspected life-threatening injuries, though, the use of WBCT or - “pan CT scan” - in trauma is an often debated topic in the emergency department (ED).

Nevertheless, WBCT is used indiscriminately in trauma cases, just on the suspicion of them being polytrauma cases [**1**] and its use is questionable in seemingly normal patients just to allow for an early emergency department (ED) discharge. Furthermore, the risk of developing Radiation-induced cancer can’t be denied, particularly in young patients exposed to high dose radiation as is the case in WBCT [**2**].

The European injury database (IDB) 2014 report stated hospital treatment of around 40 million people who had suffered accidents (in all sectors including transport, workplace and school) on an annual basis with deaths reported in 233,000 cases [**3**]. The latest diagnostic imaging dataset statistical release by the NHS, England, mentioned approximately 380 ×10^3^ total CT scan examinations performed every month in the United Kingdom, with no particular reference to polytrauma WBCT figures [**4**]. Sammy *et al*. published retrospective observational study of Trauma Audit and Research Network (TARN) data from 2012–2014 and reported WBCTs in 16.5% of polytrauma patients with more scans being done in major trauma centres than non-designated hospitals [**5**]. With the advent of major trauma centers across the UK many patients are transferred to CT after initial assessment with targets in place for this scan to be done in a relatively short time window [**6**].

For a large number of patients, there is no controversy as to whether WBCT should be included as part of their management and most centers have a Radiology pro forma with specific requesting criteria, to ensure that the examination is justified. However, there are a seemingly increasing number of patients who appear to fulfil these criteria for WBCT, and in whom the study turns out to be normal with no trauma related pathology [**7-8**].

The rapid adoption of WBCT in trauma seems to have taken place without a full examination of the risks associated with this intervention. By indiscriminately ordering WBCT, patients may have more incidental findings that lead to increased anxiety, downstream testing, and costs. A recent New York trauma registry study [**9**] found that 40% had incidental findings on their trauma CTs. Of these, 63% were considered Class 2 findings that did not require urgent evaluation, but were also not normal variants, thus requiring further work-up.

It has been reported that, 1–3% cancers worldwide are caused by medical sources of radiation. WBCT on an average exposes each patient to more than 20 mSv of effective radiation dose which increases the risk of cancer mortality of 1 in 900 with radiation dose of 24 mSv in 35-year-old male and 1 in 1,250 with radiation dose of 10–20 mSv in average 45-year adult patient [**10-12**]. Keeping these statistics in perspective, it becomes the responsibility of the medical fraternity to limit the use of whole-body CT in polytrauma patients and proper guidelines regarding the same need to be established.

In the recent years there has been a shift from indiscriminate referral to WBCT of trauma patients exposed to high energy trauma, to a more risk/benefit-oriented approach suggesting clinical prediction rules to safely omit WBCT [**13**]. Some studies point out that WBCT can safely be omitted if certain criteria are fulfilled [**14**]. Evidence based guidelines may limit radiation exposure and reduce cost in a safe way [**15**].

In our local ADAN hospital, WBCT, is an important adjunct in trauma care and it is often part of standard protocol in the initial management of polytrauma patients. In this retrospective study, we instigated an investigation to assess whether there was an overuse of WBCT in patients with blunt or penetrating trauma at our local emergency department and also to estimate the true incidence of critical injuries. We are aiming to promote a good clinical examination done pre-emptively that could prevent the need for the WBCT investigation and its undesirous effects.

## METHODOLOGY

This study was performed in the Adan Hospital, which is one the biggest general hospitals in Kuwait and covers a population of about one million and it is a tertiary care major trauma center. We carried out a retrospective database analysis of 546 polytrauma cases who referred to the radiology department, Al-Adan Hospital, for WBCT during the period of April to October 2018. WBCT requested for any other causes were excluded from this assessment. All the trauma patients were initially managed and proceed to WBCT according to the American College of Radiology (Major Blunt Trauma) [**16**].

Informed patient consent was taken during the time of the scan. All the scans were performed on a diagnostic DE CT Revolution 64 slice in helical mode with no contrast for head but IV contrast for the chest and abdomen was injected. Parameters of 120 kVp, Smart mA: (56-156), slice thickness of 5 mm, 50 cm SFOV, 0.984 pitch and BODY filter were used. We recorded age, gender, mechanism of injury, clinical requests and concerned raised, WBCT findings in regions of the cervical spine, thoracolumbar spine, chest, abdomen, pelvis and appendicular skeleton injuries (if asked for) as per our institutional reporting protocol. Dose length product (DLP) of each patient scan recorded at the end of examination.

We compared pre-test clinical requests stating the mechanism of injury and clinical query with WBCT findings and categorized the radiological findings, initially into negative and positive findings. The positive findings were further classified into major and minor injuries. The total numbers in each radiological finding were calculated and inferences discussed.

## RESULTS

A total number of 546 patients (384 male and 162 female) who were referred to the radiology to have WBCT were included in this study. In an analysis of data related to the WBCT, we found that 462 patients had been referred due to RTA (84.6%), 47 patients due to FFH (8.6%), ten patients due to burnt trauma (1.8%), 8 patients due to assault (1.5%), while the 5 patients had stable wound (0.09%), 5 patients had injury due to the fall of a heavy object (0.09%), 3 patients had a buggy injury (0.05%) and 3 patients had injury due to fallen on the back (0.05%) and 3 more patients had injury due to other traumas including blast injury, suicide and other injury lead to loss of consciousness. We noticed RTA with a 84.7 % score was the most common indication for WBCT referral. The results were presented in **Figure 1**.

**Figure 1.**
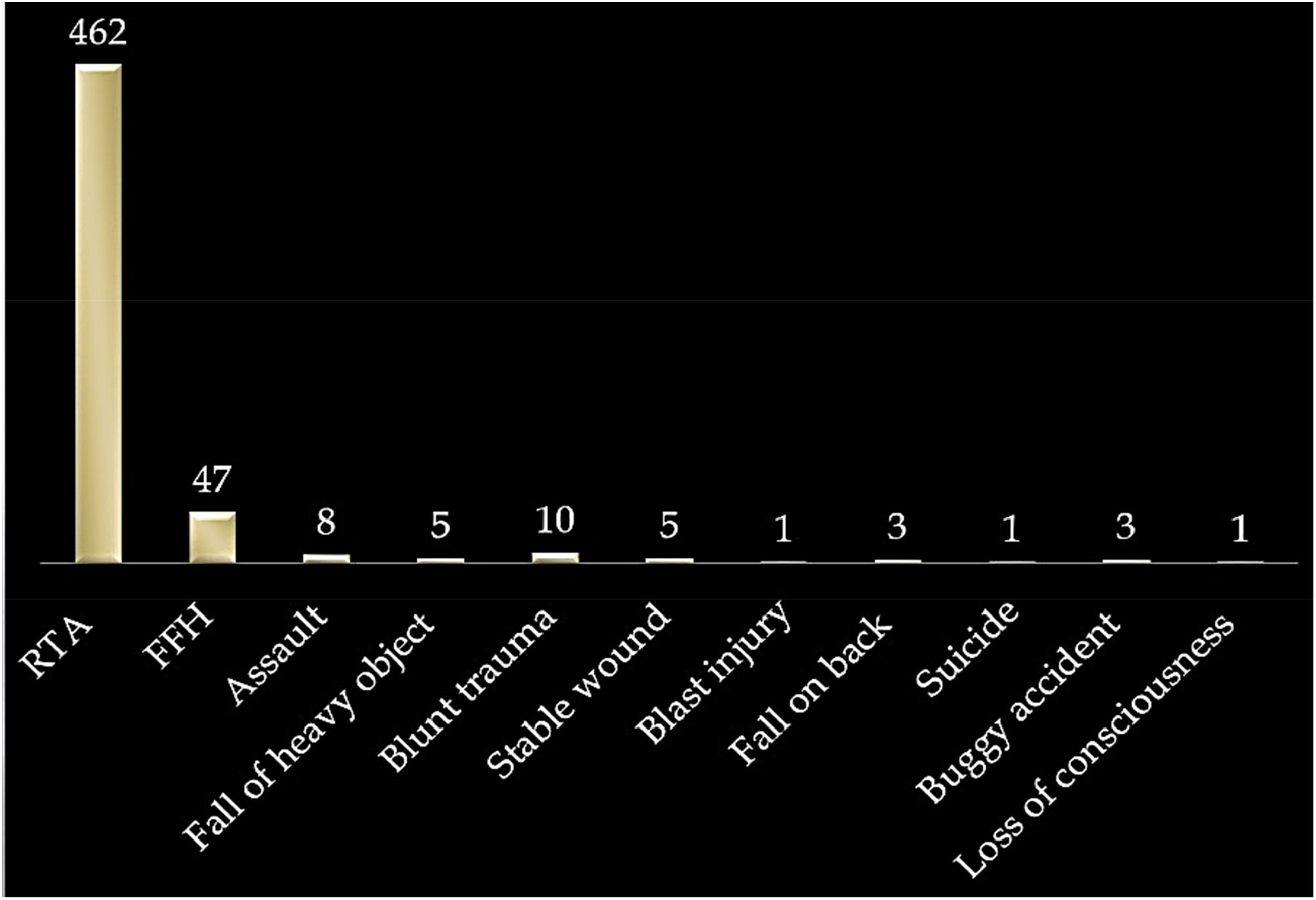
Number of patients in each subset of CT Indication.

Out of 546 patient referrals, 414 patients (75.8 %) were normal (negative finding), where 132 patients (24.2 %) had positive trauma related radiological finding of which 54 patients were found to have a major injury (9.9%) including solid organ and vertebral column injuries. Fractures were scored the highest, 75.6% of all positive finding traumas. The results were tabulated in **Figure 2**.

**Figure 2.**
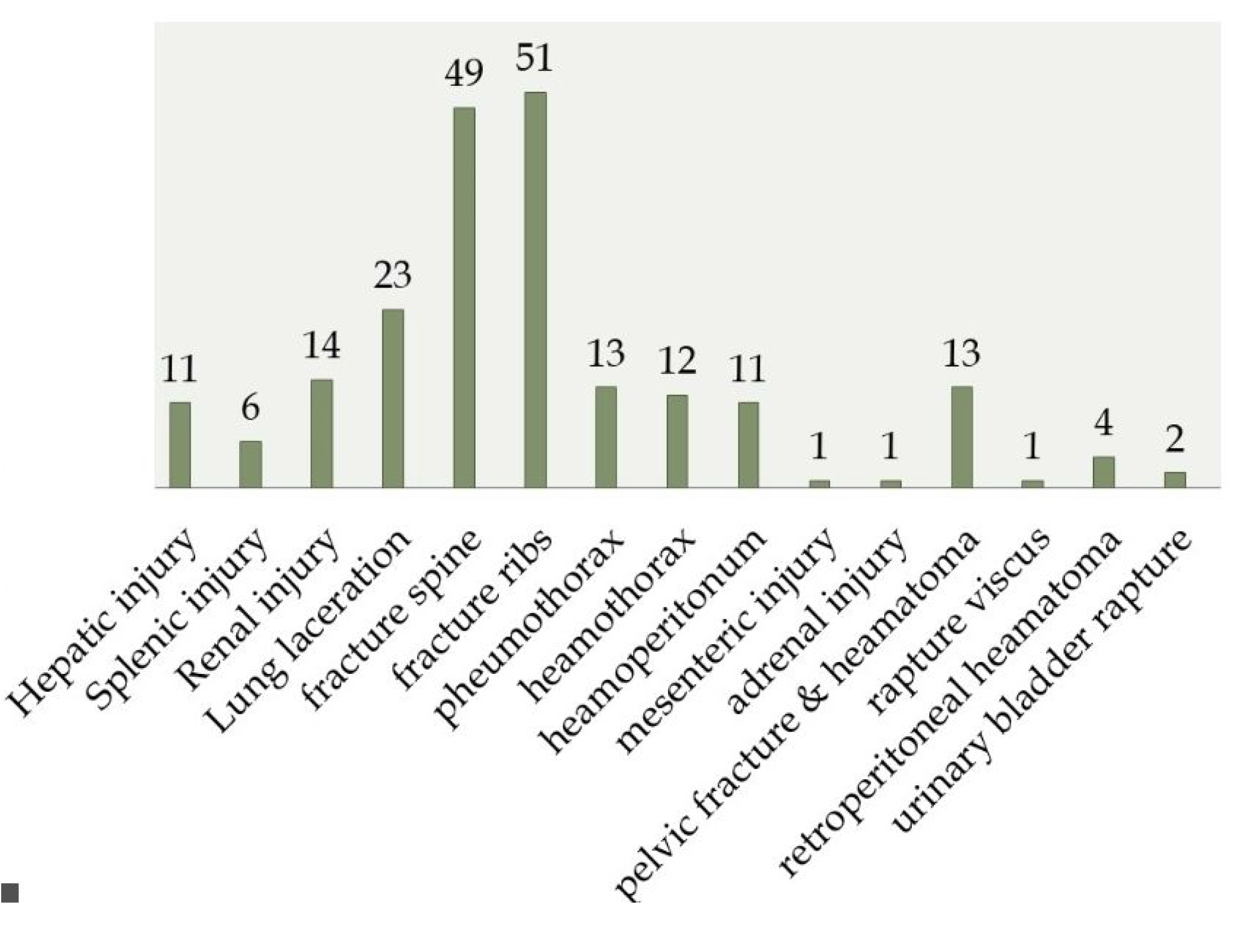
Frequency of positive patient categories based on clinical suspicion and WBCT finding.

## DISCUSSION

The use of WBCT has significantly increased in the Emergency Departments since it decreases the time needed to reach a diagnosis and decrease patient stay in emergency departments [**17**]. WBCT in polytrauma is important for planning of treatment in severely injured patients while it can be questioned in patients seemingly not injured where it is used mainly to permit early discharge from the ED. In the recent years there has been a shift from indiscriminate referral to WBCT of trauma patients exposed to high-energy trauma, to a more risk/benefit-oriented approach suggesting clinical prediction rules to safely omit WBCT [**18**]. The use of WBCT has significantly improved the management of trauma patients, though, its utilization in relation to the side effects of radiation and its financial burden are a matter for debate [**19**].

The evidence given by Huber-Wagner *et al*. [**20**] in their published paper was overwhelmingly convincing for the inclusion of whole-body CT in the pathway of multiple injured patients, as it was found beneficial in improving outcome and was therefore part of standard protocol for work-up of these patients. However, the study had several limitations and most important being preselection of high risk patients with suspicion of severe injuries, who mostly benefitted from CT scanning. On the contrary to their study, there was not a significant association found in patients suspected of major injury in our final WBCT outcome, as only 9.9% of all examined patients were suffering from a major injury. Davies *et al. a*lso proposed use of a clinical scoring system based on clinical signs of trauma to more than one body region, Glasgow Coma Scale, hemodynamic abnormality, respiratory abnormality and mechanism of injury [**21**], however, forming a clinical score was not one of the objectives of our study. There are several studies, which have also suggested the use of clinical prediction rules to safely omit unnecessary WBCTs, reducing radiation dose and cost [**22-24**].

Our findings showed a high percentage of patients were normal (74.8 %) and only 24.2 % of patients were having positive findings of which only 9.9% were found to have major injuries. RTA was the most common indication for WBCT. The results showed that we are experiencing a highly elevated use of WBCT in our hospital, considering only 24.2% of the cases were found positive and therefore it is beneficial to review the current protocol in place for utilizing WBCT in polytrauma to minimize the unnecessary requests respectively.

The use of WBCT and 24/7 availability of skilled personnel for performing and interpreting WBCT is also associated with significant cost. The cost of WBCT is equivalent to $260 at our hospital. However, this cost is negligible if the WBCT detects a significant injury, or when we make use of WBCT for admission to the intensive care unit for observation. We suggests the tabulated criteria [**Table 1**] could be used as the guide line protocol for use of WBCT in polytrauma patient, as its usage has no impact on patient care if the patient is evaluated by a trauma team to show he is mentally alert and there is only a sign of minor injuries. The risk of missing important traumatic injuries in these patients is very low.

**Table 1.** Criteria to follow for immediate WBCT

|  |  |
| --- | --- |
| **Trauma patients with the presence of one of the following vital parameters:** |  |
| o | respiratory rate >29/min or <10/min; |
| o | pulse >120/min; |
| o | systolic blood pressure < 100 mmHg; |
| o | estimated exterior blood loss > 500 ml; |
| o | Glasgow Coma Score ≤ 13; |
| o | abnormal pupillary reaction on site. |
| **OR patients with one of the following clinically suspicious diagnoses:** |  |
| o | fractures from at least two long bones; |
| o | flail chest, open chest or multiple rib fractures; |
| o | pelvic fracture; |
| o | unstable vertebral fractures; |
| o | spinal cord compression. |
| **Trauma patients not receiving total-body CT scanning:** |  |
| o | known age <18 years; |
| o | known pregnancy; |
| o | referred from another hospital; |
| o | any patient who is judged to be too unstable to undergo a CT scan and requires resuscitation or immediate operation. |

Observation of the patient and re-examination instead of WBCT imaging can be considered to minimize the radiation dose and its effects receiving by this group of often young patients that is a matter of concern at the State level. *Karla etal*. reported in their publication that in the USA, two thirds of the radiation received from imaging is related to CT scan and this has elevated the risk of radiation mortality to 12.5 cases per 10000 CT scan of population [**25**]. The risk of developing radiation induced cancer is significant for young people if they exposed to relatively high dose of radiation as in case of WBCT [**Figures 3-4**].

**Figure 3.**
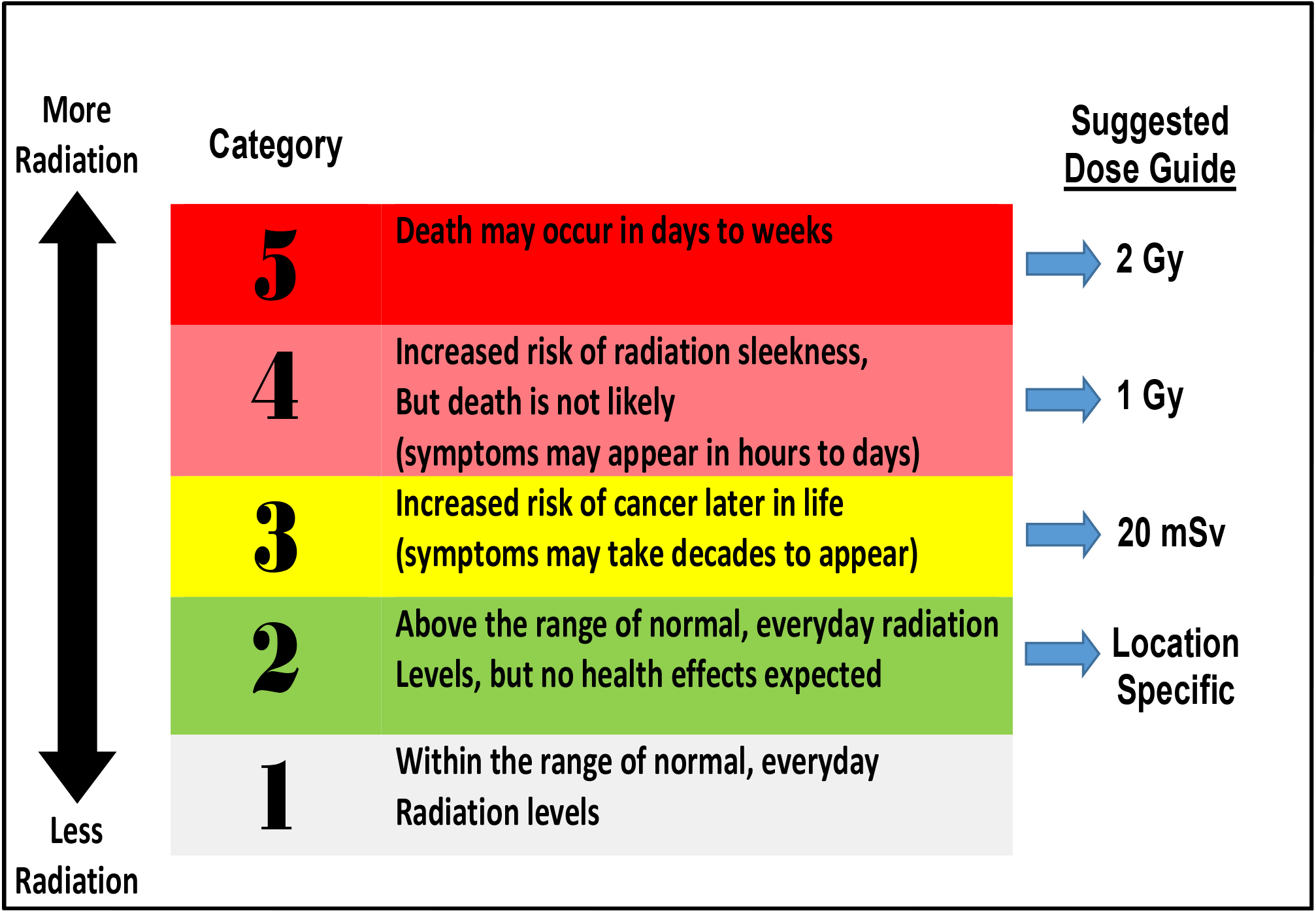
Radiation health effects in relation to radiation dose

**Figure 4.**
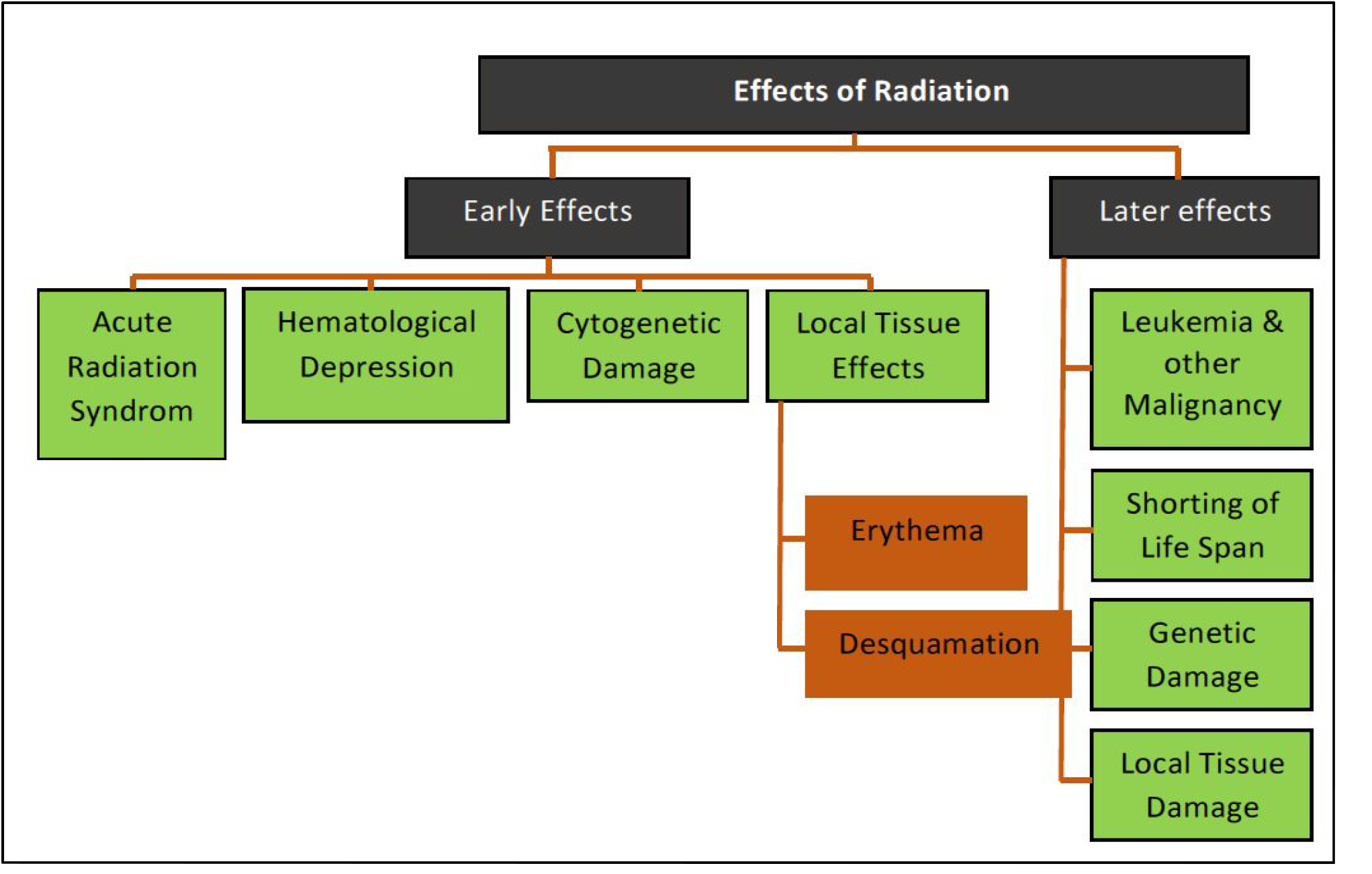
Early and long term effect of radiation in relation to radiation dose.

## CONCLUSIONS

The results of our retrospective study revealed that there is an overuse of WBCT in trauma patients in the emergency and surgical departments in our hospital as low incidence of significant injuries (9.9%) does not justify the routine use of WBCT for every trauma patient which in turn leads to poor utilization of radiology department resources and having an impact on patient safety and wellbeing. Evidence based guidelines may limit radiation exposure and reduce cost in a safe way. We share our experience on this controversial topic and sincerely hope that with an increase in the number of published studies on this topic, proper guidelines for use of WBCT in early management of polytrauma patients can be formulated.

## Data Availability

all relevant data stated in the manuscript are available in request at any time.

## ACKNOWLEDGMENT

The authors would like to thank Hany Elrahman, department of medical imaging, for assisting with CT technical data and collecting the related references.

